# An investigation into ethnic group and deprivation inequalities in influenza and COVID-19 hospitalizations, post-pandemic

**DOI:** 10.1101/2025.08.14.25333430

**Authors:** Meg Scott, Lucy Browning, Tirion Roberts, Jessica De Boer, Zoe A. Richardson, Leonardo Gada

## Abstract

Health system decision-makers need evidence on how modifiable factors contribute to inequalities to inform effective interventions. We examined ethnic group and deprivation inequalities in influenza and COVID-19 hospitalizations in England (2023 – 2024) among vaccine-eligible individuals, using causal mediation analysis to assess the roles of deprivation, vaccination uptake and comorbidities.

For influenza, hospitalization odds were highest among Pakistani and Bangladeshi groups, compared with White British individuals (OR 2.0, 95% CI: 1.83, 2.10; OR 1.6, 95% CI: 1.39, 1.80, respectively). Deprivation mediated 26% of the total ethnic group effect for the Pakistani group and 41% for the Bangladeshi group, whilst comorbidities contributed little. Ethnic group differences in COVID-19 hospitalizations were minimal in this post-pandemic period.

For both infections, the most deprived group had disproportionately high hospitalization compared to the least deprived (influenza OR 1.7, 95% CI: 1.63, 1.79; COVID-19 OR 1.5, 95% CI: 1.43, 1.55. For influenza, vaccination uptake mediated 17-26% of deprivation effects, whilst for COVID-19, there were no meaningful residual deprivation inequalities after including vaccination uptake and comorbidities.

Vaccination uptake and deprivation mediate a substantial proportion of inequalities in hospitalization for influenza and COVID-19 in the vaccine-eligible cohort. Improving vaccination coverage in under-vaccinated groups should remain a public health priority.

## Introduction

Health inequalities are unfair, avoidable and systematic differences in morbidity and mortality between population groups. Reducing inequalities is a core aim of health policy. To target interventions effectively, decision-makers require timely, comparable measures of inequalities and evidence on the relative contribution of modifiable factors. This is challenging without detailed data, and improving evidence on ethnic group and deprivation inequalities remains a key priority for health protection in England [1].

Influenza and COVID-19 contribute substantially to morbidity and mortality in England, with unequally distributed burdens across ethnic groups and deprivation levels. This includes inequalities for infection itself, and for severe outcomes following infection – hospitalization, intensive care admissions and mortality [2–6]. Deprivation itself is also a well-established risk factor for influenza and COVID-19 [7]. Both infections have established vaccination programmes, with wide coverage but notable inequalities in uptake [8, 9]. However, vaccination inequalities are often monitored separately from inequalities in hospitalization and comorbidities, limiting understanding of how these factors interact to produce unequal outcomes.

Recent UK Health Security Agency analyses [7] found significant inequalities in hospitalization for influenza and COVID-19, by deprivation and ethnic group in England (winter 2022 to 2023). Influenza admission rates for the most deprived areas were 2.6 times higher than the least deprived areas, and 2.1 times higher for COVID-19. For influenza (but not COVID-19) there were persistent differences in hospitalization rates between ethnic groups, for instance, the Pakistani group had influenza hospitalization rates 2.7 times higher than the White group. Post-pandemic, ethnic group inequalities for COVID-19 have been comparably lower than during the pandemic [10].

Vaccination reduces infection risk and disease severity, but uptake remains consistently lowest in the most deprived areas, and among some ethnic groups. In reporting closest to the time period of this study (2023 to 2024), 79% of those aged ≥ 65 from the least deprived areas were vaccinated for COVID-19 in the 6 months prior, compared to 59% in the most deprived areas [8]. Influenza vaccine uptake was also lower in the most deprived areas compared to the least deprived areas, across all vaccination cohorts [8]. There were considerable ethnic group inequalities in vaccine uptake; for COVID-19, vaccine uptake was lowest for the Pakistani (18%), and Bangladeshi groups (24%), and highest for the White British (76%) and White Irish groups (64%) [8]. For influenza, vaccine uptake was lowest for the Black or Black British Caribbean (46%) and the Black or Black British African groups (48%), and highest for the White British (82%) and White Irish groups (76%) [8].

Comorbidities which predispose individuals to a higher risk from COVID-19 and influenza are detailed in the clinical risk groups in the Green Book, which provides information for public health professionals on immunization [11]. Risks for many of these conditions are higher among certain ethnic groups, including cardiovascular disease, diabetes and multimorbidity [12–14]. People from Indian, Pakistani, Bangladeshi, Black African, Black Caribbean and people belonging to “Any other Black ethnic group”, “Any other Asian ethnic group”, or mixed ethnic groups may also have higher risk of developing multiple long-term conditions [15]. Comorbidities are also highest for those living in deprived areas, with large differences by deprivation for mortality involving lung cancer and asthma, two risk factors for COVID-19 and influenza [12–14]. These conditions form part of the eligibility criteria for vaccination programmes, reflecting existing policy efforts to mitigate severe outcomes among higher-risk groups.

Previous studies have shown that comorbidities, vaccination coverage and socioeconomic, demographic and geographic factors together account for differences in COVID-19 mortality across ethnic groups during the pandemic [16]. However, evidence on the relative importance of specific mediators remains limited, especially for influenza and for COVID-19, post-pandemic. In this paper, we use causal mediation analysis with linked administrative data on hospitalizations, vaccination, and laboratory testing, to estimate the extent to which deprivation, vaccination uptake and comorbidities explain ethnic group and deprivation inequalities in influenza and COVID-19 hospitalizations, 1 September 2023 – 31 March 2024, for the vaccine-eligible cohort.

## Methods

### Study population

The study population was defined using age and clinical risk data within the Immunisation Information System (IIS), based on the Joint Committee on Vaccination and Immunisation criteria for vaccine eligible cohorts. For influenza and COVID-19 during winter 2023 – 2024, eligibility included individuals aged ≥ 65 years, those in clinical risk groups (Supplementary Materials), care home residents, household contacts of immunosuppressed individuals, and frontline health and social care workers. For influenza only, children aged 2-16 years were also eligible.

Analyses were restricted to vaccine-eligible individuals to assess the contribution of vaccination uptake to hospitalization risk, ensuring observed differences were not due to eligibility criteria. All analyses were repeated for the whole population as sensitivity analyses (Supplementary Tables 9 – 13). Results presented in the main paper relate to the vaccine-eligible cohort.

### Data sources

Hospitalizations between 1 September 2023 and 31 March 2024 were identified from the Hospital Episode Statistics Admitted Patient Care dataset [17], restricted to one observation per individual. Vaccination data were obtained from IIS, which captures all vaccination events for individuals registered with a general practitioner in England, include vaccinations delivered in pharmacies, schools, and other settings. The number of individuals without an NHS number has been assessed to be small, but is likely to disproportionately include more vulnerable populations, such as migrants, asylum seekers and undocumented individuals [18]. As a result, whilst it is unlikely that vaccination events are missed from the data, vaccination inequalities may be underestimated if unvaccinated individuals from these groups are excluded from the denominator. Overall, IIS vaccination records and denominators are the most complete record available in England and have been assessed as highly accurate, as compared to enhanced surveillance from the COVID-19 pandemic [18]. All vaccination events between 1 September 2023 and 31 March 2024 were included; the earliest cohort commencement, was 1 September 2023 for children’s influenza vaccinations.

Laboratory-confirmed COVID-19 and influenza tests were sourced from the Second Generation Surveillance System, and linked to hospitalizations. Data were linked using unique NHS numbers and accessed in accordance with NHS Digital Data Sharing Agreements on a secure platform. Analyses were conducted on aggregated subgroup-level data, ensuring no individual was identifiable.

### Outcomes, exclusions and covariates

Hospitalization with influenza or COVID-19 was the primary outcome. Variable definitions are provided in Supplementary Table 5. Admissions without diagnostic or laboratory confirmation, or with missing covariates were excluded (5% of influenza total eligible cohort, 3% of COVID-19 eligible cohort, Supplementary Table 4)

### Statistical analyses and causal framework

Directed acyclic graphs (Figure 1) were used to define causal structures and adjustment sets [19]. Ethnic group and deprivation were analysed separately as exposures. When ethnic group was the exposure, age, sex and region were treated as competing exposures, deprivation, comorbidities and vaccination uptake were treated as mediators, with deprivation and comorbidities as intermediate confounders. Total causal effects were estimated using models that included competing exposures, as their inclusion improved model fit (Supplementary Table 6). When deprivation was the exposure, ethnic group, age, sex and region were pre-exposure confounders, and comorbidities and vaccination uptake were mediators, with comorbidities as an intermediate confounder.

**Figure 1:**
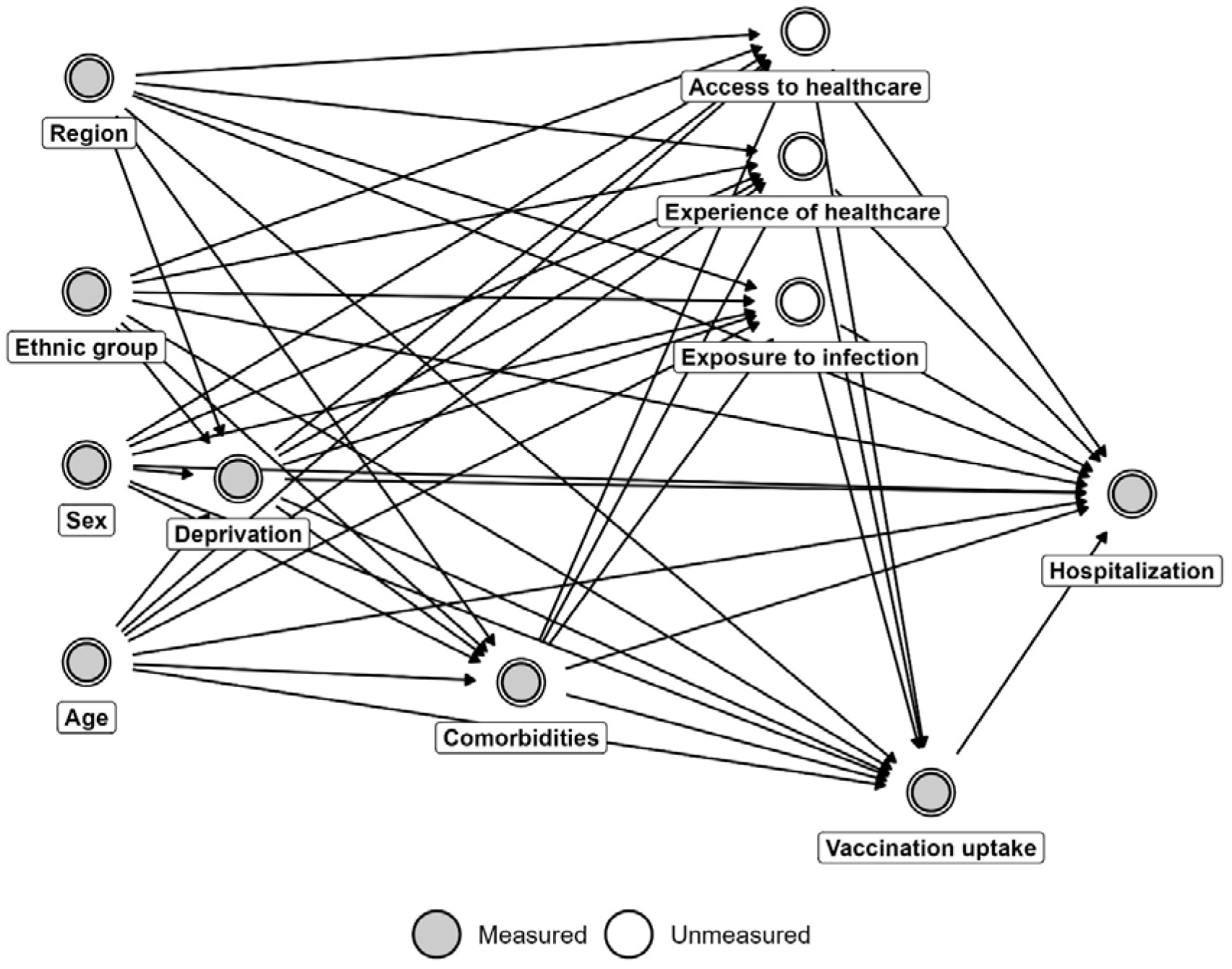
Directed acyclic graph (DAG) for the effect of ethnic group and deprivation on the outcome of hospitalization with influenza or COVID-19.

Unmeasured mediators (exposure to infection, experience of healthcare, and access to healthcare) were assumed to crystallize at the same time point in the context of this analysis and are considered a limitation of results.

The DAG was constructed following recommended practice for causal inference, with variables arranged in temporal order from left to right and arcs drawn using forward saturation [19]. Assumed relationships were informed by Office for National Statistics Census (ONS) 2021 population statistics and sociodemographic cross-tabulations [20], together with existing literature on comorbidities, vaccination uptake, access to healthcare, experience of healthcare, and infection risk (references 3–18).

### Models and mediation analysis

Hospitalizations were modelled using weighted binomial regression to estimate total causal effects (equations in Supplementary Table 16). Mediation analyses were conducted using regression-based methods when no intermediate confounding was present [21] and g-formula methods when intermediate confounders were included [22] [23]. Estimation of mediated effects was limited to focal relationships with, at most, one intermediate confounder, as the R package used here cannot accommodate multiple sequential mediators. Therefore, for ethnic group as exposure, we estimated mediation by deprivation, and mediation by comorbidities, conditioning on deprivation, but we were unable to estimate mediation by vaccination uptake. For deprivation as exposure, we estimated mediation by comorbidities, and then separately estimated mediation by vaccination uptake, while conditioning on comorbidities. Comorbidities and vaccination uptake mediator or intermediate confounder models were estimated using weighted binomial models, deprivation models were estimated using weighted multinomial models. All models were fitted using the *cmest* function in *CMAverse* (R version 4.3.2) [23].

### Interpretation

Effects were reported as odds ratios (ORs). ORs (and associated 95% confidence intervals) exceeding 1.25 or below 0.80 were interpreted as disproportionate, following Race Equality Unit guidance [24]. ORs were interpreted as statistically significant when associated 95% confidence intervals did not overlap with 1.0 (parity). Mediation results were presented only for ethnic groups or deprivation levels where total effects indicated disproportionality. Given the rarity of hospitalization (0.07% of influenza total eligible cohort, 0.14% COVID-19 eligible cohort), ORs were considered meaningful approximations to risk ratios. Proportion mediated was estimated using differences in predicted risks on the risk scale and therefore does not correspond precisely to the ratio of odds ratios.

## Results

### Descriptive analysis

The eligible vaccination cohort included 28.4 million individuals for influenza and 19.1 million for COVID-19, with 19,077 influenza hospitalizations and 27,361 COVID-19 hospitalizations between 1 September 2023 and 31 March 2024. For influenza, substantial differences between ethnic groups (compared to the White British group) and deprivation levels (compared to the least deprived group) were observed in hospitalization, vaccination uptake and comorbidities. For COVID-19, substantial differences were present for vaccination uptake and comorbidities, but hospitalization differences by ethnic group were smaller (Tables 1 – 2).

### Influenza

#### Total causal effects

In the adjusted total causal effect model for ethnic group, including age, sex and region as competing exposures (model 1, Table 3a), the Pakistani and Bangladeshi groups had disproportionately high odds of influenza hospitalization compared with White British individuals (OR 2.0, 95% CI: 1.83, 2.10 and OR 1.6, 95% CI: 1.39, 1.80 respectively). No ethnic group had disproportionately low odds of influenza hospitalization.

In the adjusted total causal effect model for deprivation (model 2, Table 4a), adjusting for ethnic group, age, sex and region, as pre-exposure confounders, individuals in the more deprived areas (IMD1, IMD2) had disproportionately high odds of influenza hospitalization, compared to the least deprived areas. The most deprived area (IMD1) had odds 1.7 (95% CI: 1.64, 1.79) times higher than the least deprived.

**Table 1:**
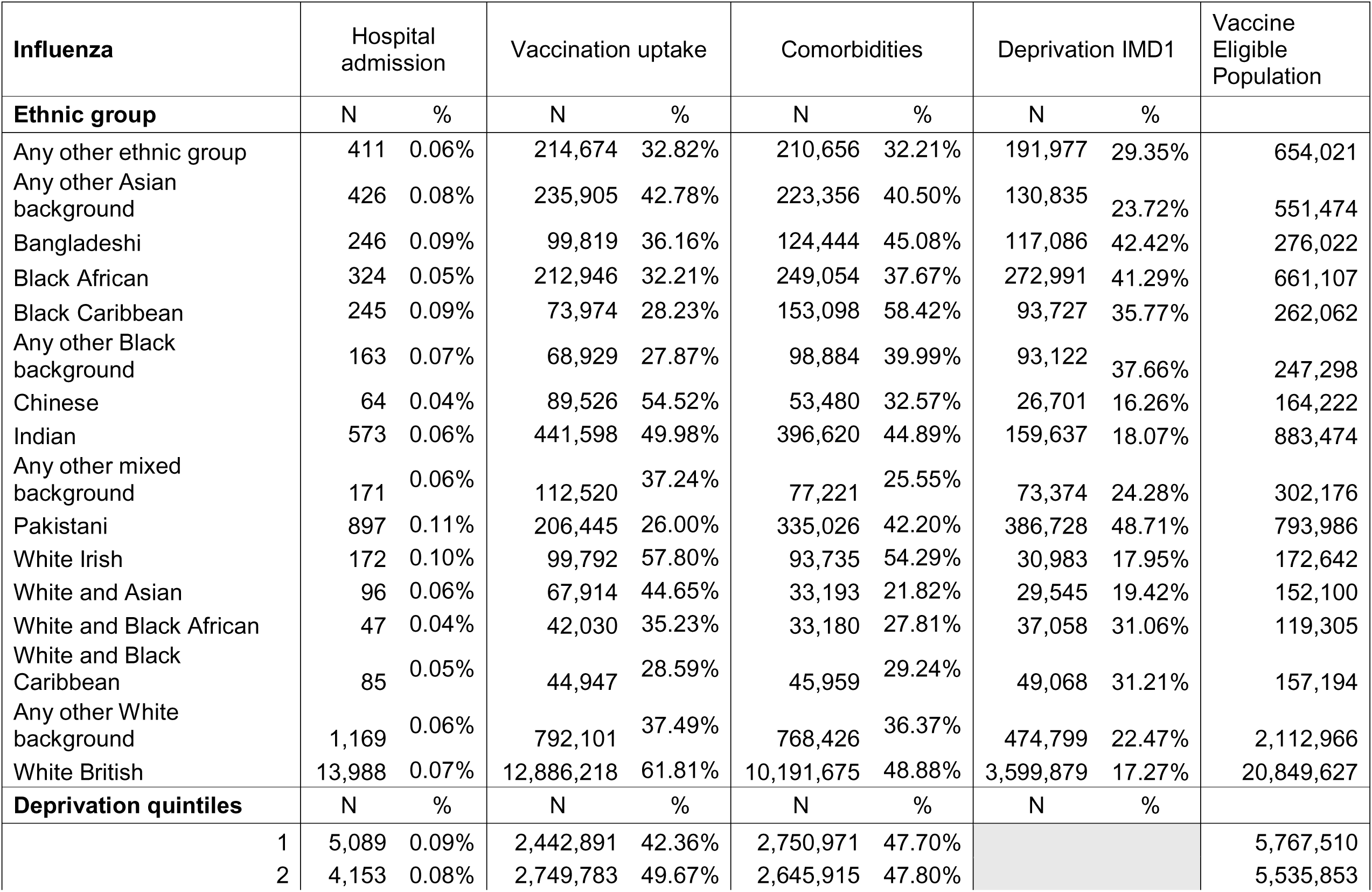
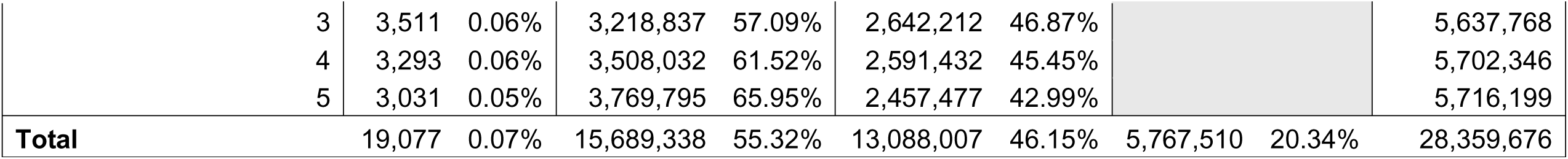
Descriptive characteristics of the influenza vaccine-eligible cohort.

**Table 2:**
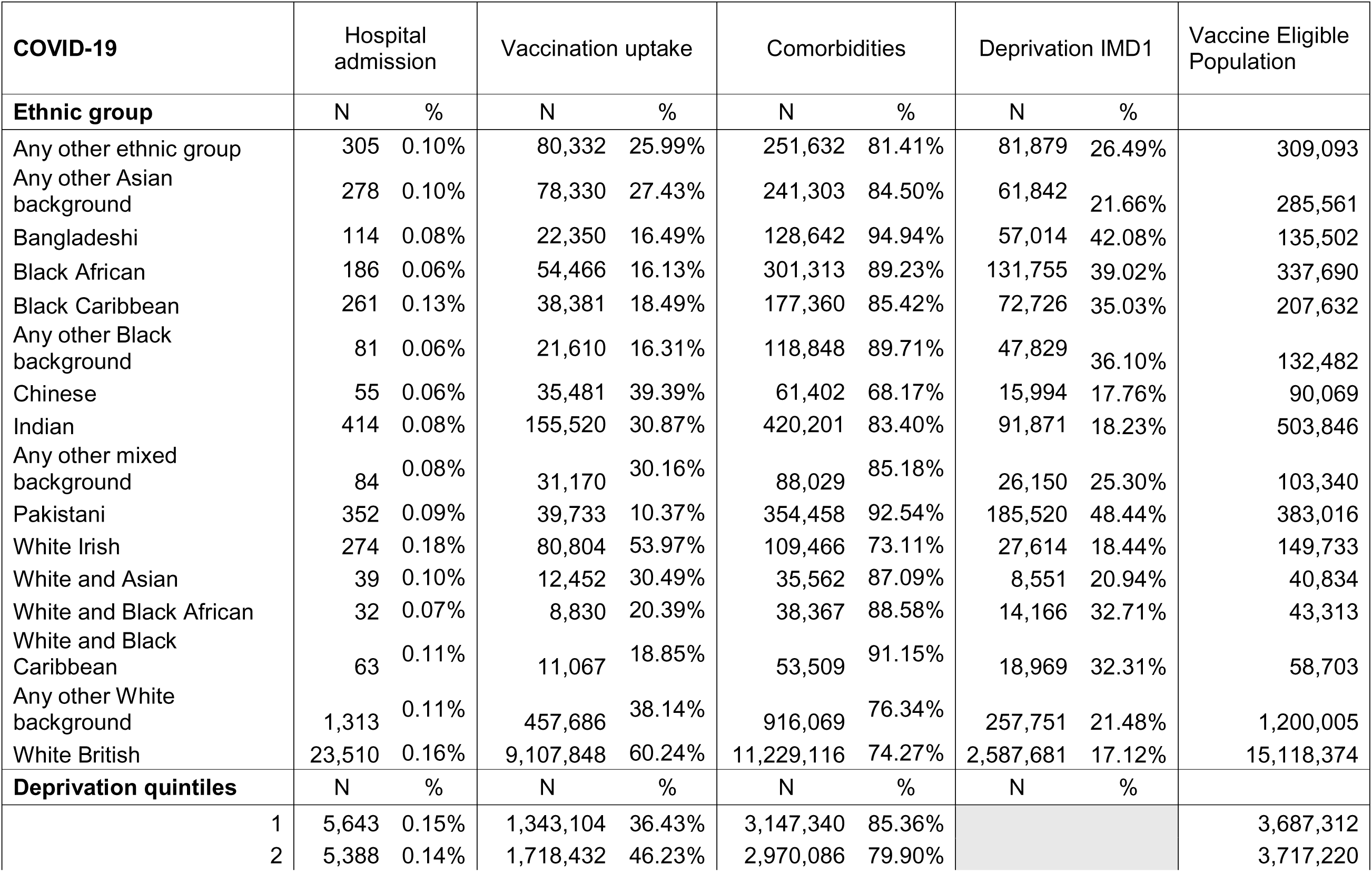
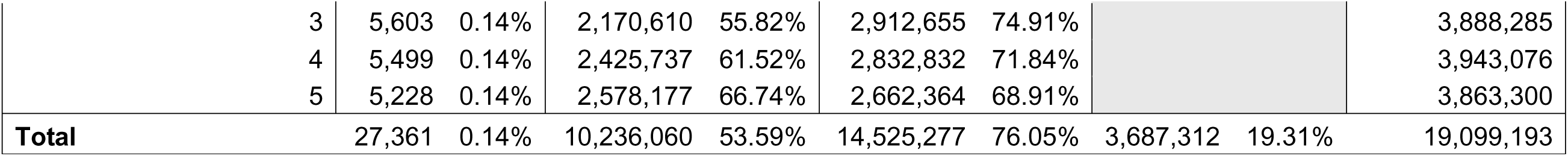
Descriptive characteristics of the COVID-19 vaccine-eligible cohort.

**Table 3a:**
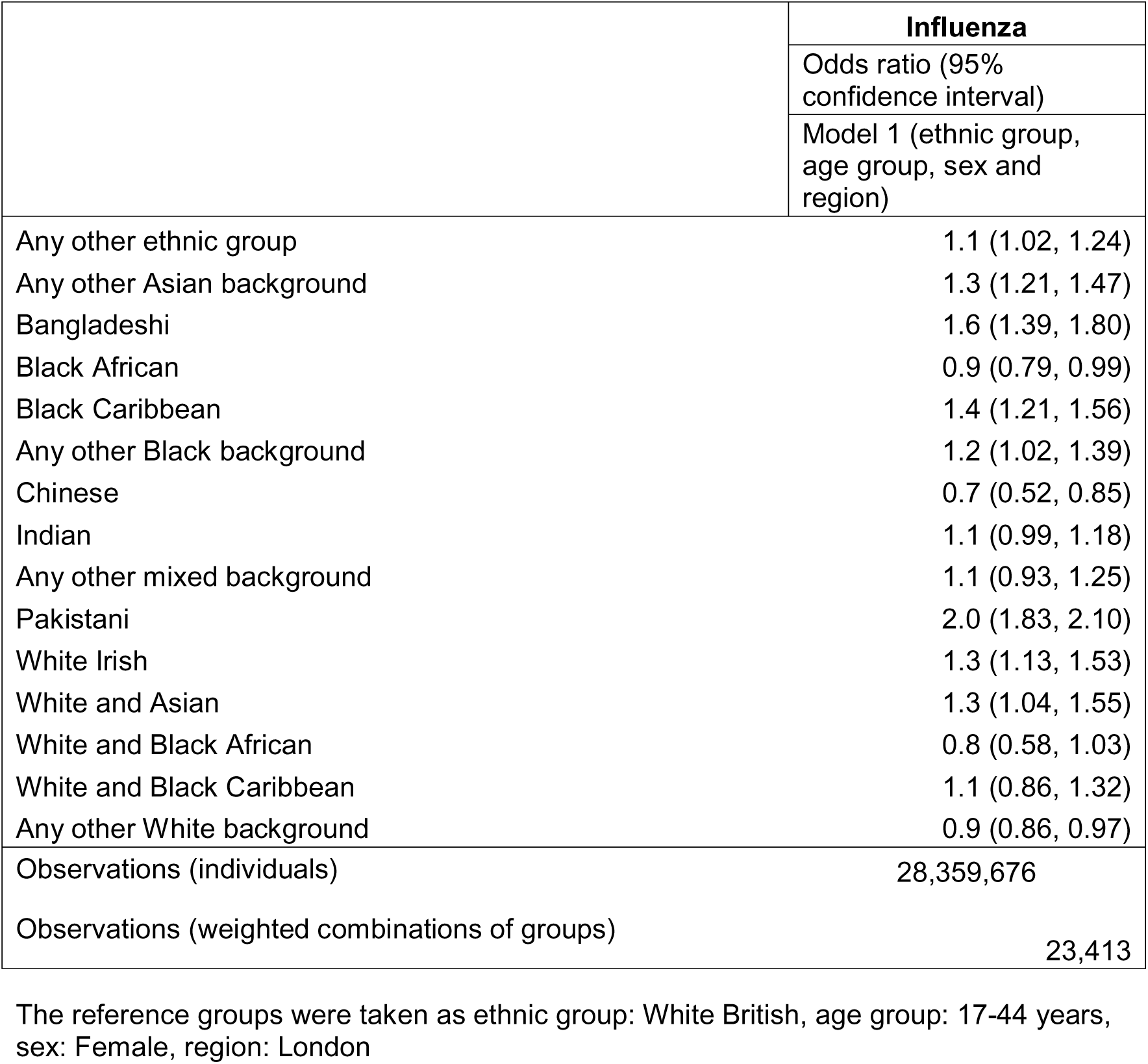
Adjusted odds ratios (95% confidence intervals) for influenza by ethnic group, Model 1.

**Table 3b:** Adjusted odds ratios (95% confidence intervals) for COVID-19 by ethnic group, Model 1.

|  | **COVID-19** |
| --- | --- |
|  | Odds ratio (95% confidence interval) |
|  | Model 1 (ethnic group, age group, sex and region) |
| Any other ethnic group | 0.8 (0.69, 0.86) |
| Any other Asian background | 0.8 (0.68, 0.86) |
| Bangladeshi | 0.8 (0.65, 0.95) |
| Black African | 0.5 (0.44, 0.58) |
| Black Caribbean | 0.9 (0.79, 1.01) |
| Any other Black background | 0.6 (0.45, 0.70) |
| Chinese | 0.4 (0.32, 0.55) |
| Indian | 0.6 (0.53, 0.65) |
| Any other mixed background | 0.7 (0.55, 0.85) |
| Pakistani | 0.8 (0.73, 0.91) |
| White Irish | 1.1 (0.97, 1.23) |
| White and Asian | 0.8 (0.62, 1.16) |
| White and Black African | 0.7 (0.49, 0.98) |
| White and Black Caribbean | 1.0 (0.80, 1.31) |
| Any other White background | 0.8 (0.74, 0.83) |
| Observations (individuals) | 19,099,193 |
| Observations (weighted combinations of groups) | 16,722 |

The reference groups were taken as ethnic group: White British, age group: 17-44 years, sex: Female, region: London

**Table 4a:** Adjusted odds ratios (95% confidence intervals) for influenza by deprivation (IMD quintiles), Model 2.

|  | **Influenza** |
| --- | --- |
|  | Odds ratio (95% confidence interval) |
|  | Model 2 (ethnic group, age group, sex, region and deprivation) |
| IMD - 1 (most deprived) | 1.7 (1.63, 1.79) |
| IMD - 2 | 1.4 (1.36, 1.50) |
| IMD - 3 | 1.2 (1.12, 1.24) |
| IMD - 4 | 1.1 (1.03, 1.13) |
| Observations (individuals) | 28,359,676 |
| Observations (weighted combinations) | 23,413 |

**Table 4b:**
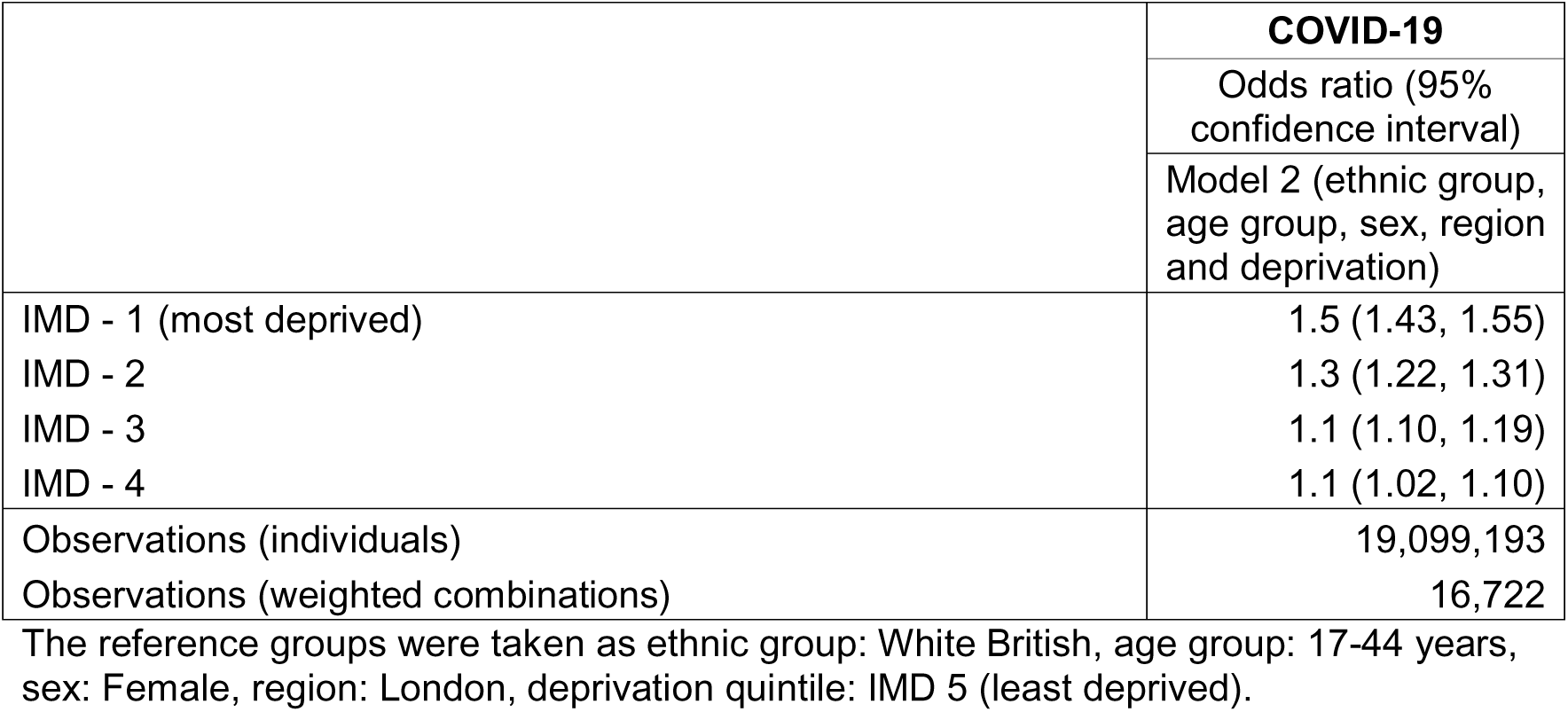
Adjusted odds ratios (95% confidence intervals) for COVID-19 by deprivation (IMD quintiles), Model 2.

### Mediation analysis

#### Ethnic group as exposure

Mediation analyses were conducted for Pakistani and Bangladeshi groups only, as the only ethnic groups with disproportionately high odds of hospitalisation (Table 3a). For both groups, indirect effects through deprivation were statistically significant. The indirect effect OR was 1.1 (95% CI: 1.08, 1.19) for the Pakistani group and 1.2 (95% CI: 1.10, 1.23) for the Bangladeshi group (Figure 2, Table 5a), corresponding to 26% and 41% of the total causal effect, respectively. Indirect effects through comorbidities, conditioning on deprivation, were not significant for either ethnic group (Table 5b).

**Figure 2:**
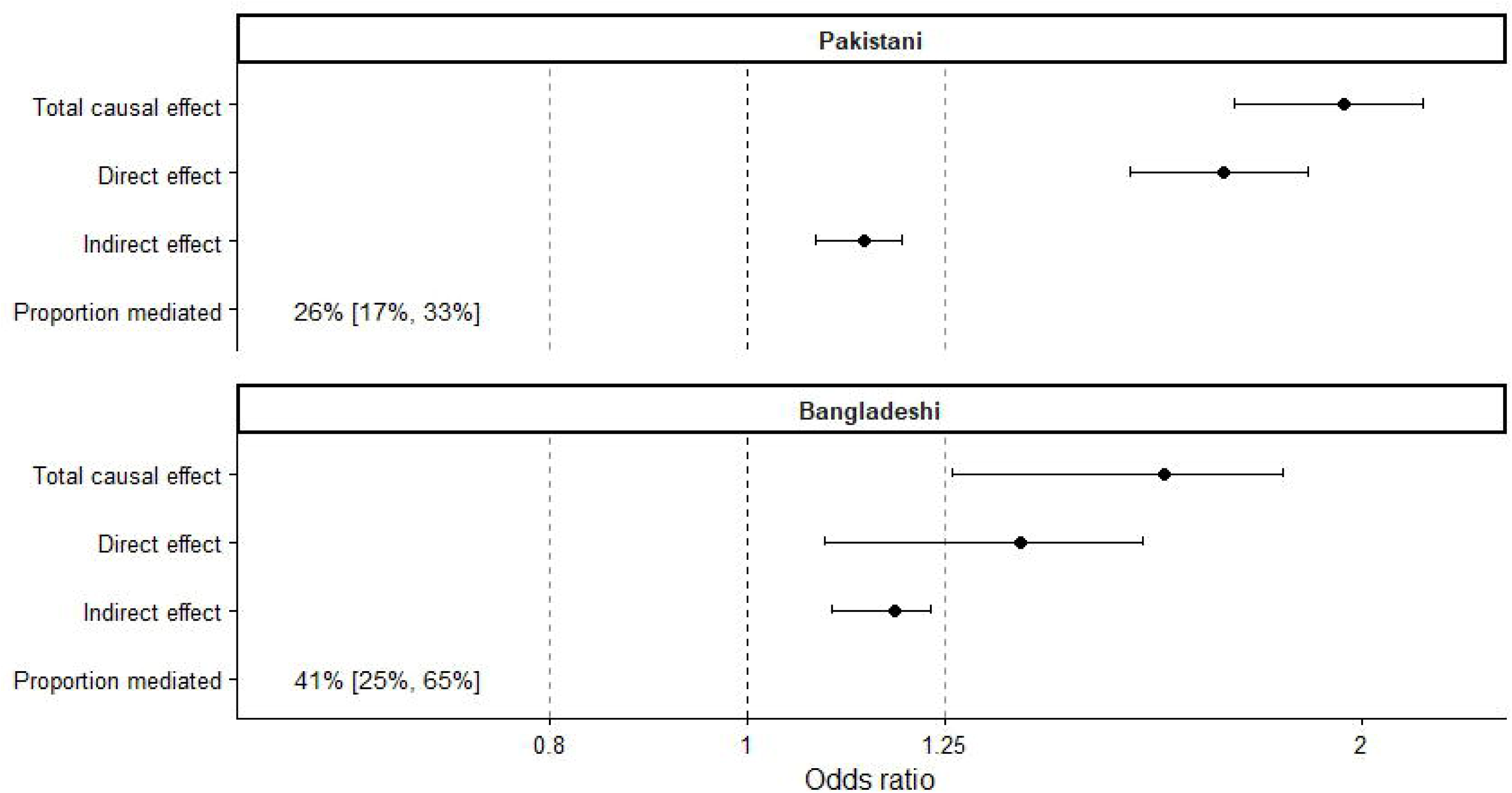
Effect of ethnic group on influenza hospitalization with mediation by deprivation, vaccine-eligible cohort, England, winter 2023 to 2024

**Table 5a:** Effect of ethnic group on influenza hospitalization, proportion mediated by deprivation.

|  | Influenza |  |  |  |
| --- | --- | --- | --- | --- |
|  | Total causal effect | Direct effect | Indirect effect | Proportion mediated |
| Pakistani | 2.0 (1.73, 2.14) | 1.7 (1.54, 1.88) | 1.1 (1.08, 1.19) | 0.26 (0.17, 0.33) |
| Bangladeshi | 1.6 (1.26, 1.83) | 1.4 (1.09, 1.56) | 1.2 (1.10, 1.23) | 0.41 (0.25, 0.65) |

The reference group was taken as ethnic group: White British
Exposure: ethnic group; Competing exposures: age group, sex, region; Mediator: deprivation

**Table 5b:** Effect of ethnic group on influenza hospitalisation, proportion mediated by comorbidities, conditioning on deprivation.

|  | Influenza |  |  |  |
| --- | --- | --- | --- | --- |
|  | Total causal effect | Direct effect | Indirect effect | Proportion mediated |
| Pakistani | 2.0 (1.73, 2.13) | 1.9 (1.69, 2.00) | 1.1 (0.98, 1.11) | 0.14 (-0.04, 0.20) |
| Bangladeshi | 1.5 (1.28, 1.89) | 1.5 (1.23, 1.78) | 1.0 (1.00, 1.12) | -0.10 (-0.01, 0.30) |

The reference group was taken as ethnic group: White British
Exposure: ethnic group; Competing exposures: age group, sex, region; Mediator: comorbidities; Intermediate confounder: deprivation

The direct effect remained statistically significant for both ethnic groups after accounting for deprivation, and remained disproportionate for the Pakistani group, therefore pathways beyond deprivation likely contributed to the excess risk. Part of both the indirect effect and direct effect could occur through vaccination uptake, as we did not decompose this factor in this analysis, or through other unmeasured factors.

### Deprivation as exposure

Mediation analyses were performed for IMD1 and IMD2 only. For individuals in the most deprived area (IMD1), the indirect effect mediated by vaccination uptake, conditioning on comorbidities, was significant (Figure 3, Table 6a) (OR 1.1; 95% CI: 1.02, 1.13), accounting for 17% of the total causal effect. A similar pattern was observed for IMD2, (indirect OR 1.1; 95% CI: 1.00, 1.12; proportion mediated 26%). Indirect effects through comorbidities alone were not significant (Table 6c). Direct effects of deprivation were statistically significant for both IMD1 and IMD2, and remained disproportionate for IMD1, after accounting for vaccination uptake, indicating incomplete mediation.

**Figure 3:**
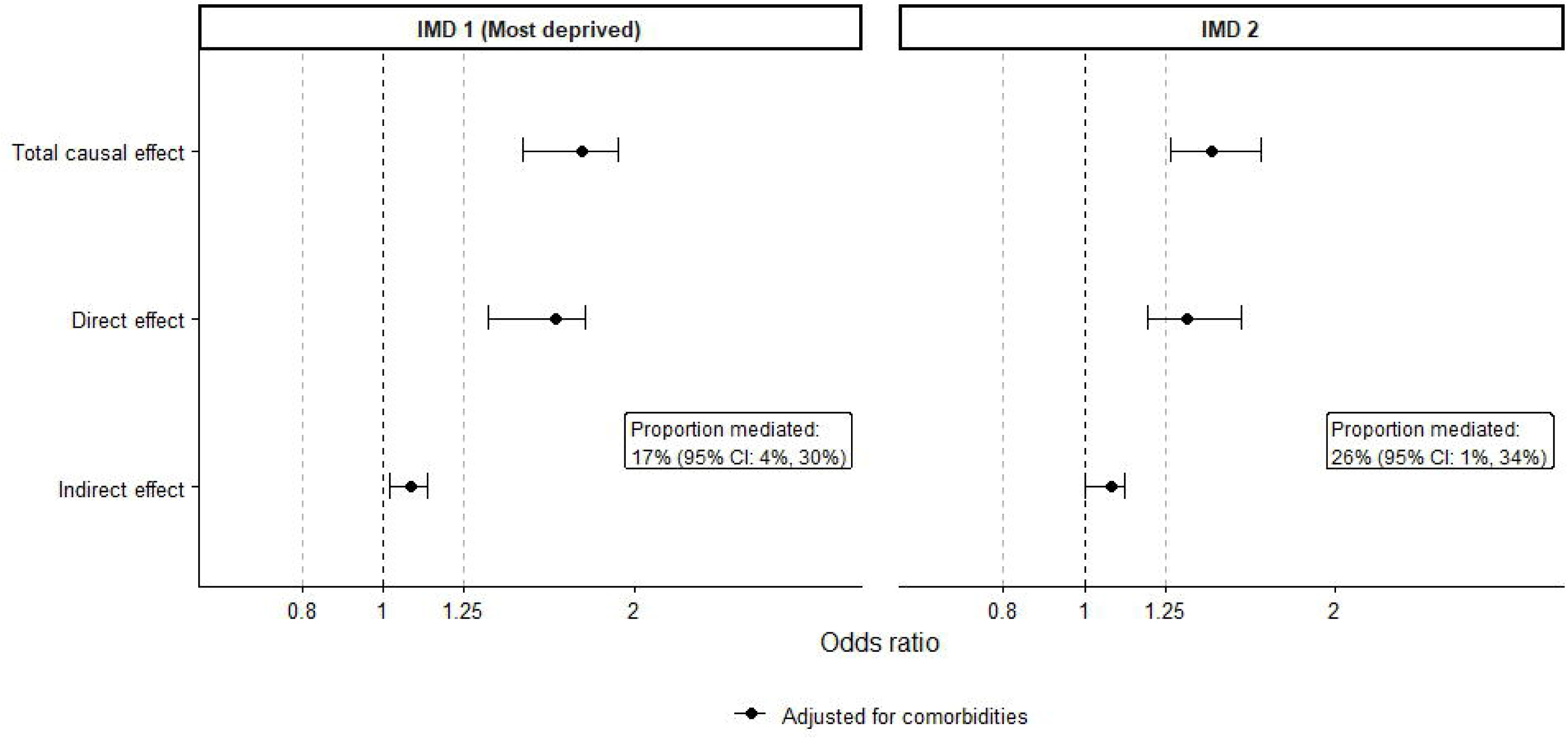
Effect of deprivation on influenza hospitalization with mediation by vaccination status, vaccine-eligible population, England, winter 2023 to 2024

**Table 6a:** Effect of deprivation on influenza hospitalization, mediation by vaccination uptake, conditioning on comorbidities.

|  | Influenza |  |  |  |
| --- | --- | --- | --- | --- |
|  | Total causal effect | Direct effect | Indirect effect | Proportion mediated |
| IMD - 1 | 1.7 (1.47, 1.91) | 1.6 (1.34, 1.75) | 1.1 (1.02, 1.13) | 0.17 (0.04, 0.30) |
| IMD - 2 | 1.4 (1.27, 1.63) | 1.3 (1.19, 1.54) | 1.1 (1.00, 1.12) | 0.26 (0.01, 0.34) |

The reference group was taken as deprivation quintile: IMD 5 (least deprived). Only IMD quintile pairwise comparisons with disproportionate total causal effect were included in the mediation analysis (IMD1, IMD2)
Exposure: deprivation; Pre-exposure confounders: age group, sex, region, ethnic group; Mediator: vaccination status; Post-exposure confounder: comorbidities

**Table 6b:** Effect of deprivation on COVID-19 hospitalization, mediation by vaccination uptake, conditioning on comorbidities.

|  | COVID-19 |  |  |  |
| --- | --- | --- | --- | --- |
|  | Total causal effect | Direct effect | Indirect effect | Proportion mediated |
| IMD - 1 | 1.2 (0.78, 3.68) | 1.0 (0.72, 3.15) | 1.2 (1.01, 1.20) | 0.97 (-0.74, 2.00) |

The reference group was taken as deprivation quintile: IMD 5 (least deprived). Only IMD quintile pairwise comparisons with disproportionate total causal effect were included in the mediation analysis (IMD1)
Exposure: deprivation; Pre-exposure confounders: age group, sex, region, ethnic group; Mediator: vaccination status; Post-exposure confounder: comorbidities

**Table 6c:** Effect of deprivation on influenza hospitalization, mediation by comorbidities.

|  | Influenza |  |  |  |
| --- | --- | --- | --- | --- |
|  | Total causal effect | Direct effect | Indirect effect | Proportion mediated |
| IMD - 1 | 1.7 (1.51, 1.90) | 1.6 (1.48, 1.75) | 1.1 (0.95, 1.17) | 0.18 (-0.14, 0.32) |
| IMD - 2 | 1.4 (1.24, 1.58) | 1.4 (1.24, 1.46) | 1.0 (0.94, 1.14) | 0.14 (-0.29, 0.39) |

The reference group was taken as deprivation quintile: IMD 5 (least deprived). Only IMD quintile pairwise comparisons with disproportionate total causal effect were included in the mediation analysis (IMD1, IMD2)
Exposure: deprivation; Pre-exposure confounders: age group, sex, region, ethnic group; Mediator: comorbidities

**Table 6d:** Effect of deprivation on COVID-19 hospitalization, mediation by comorbidities.

|  | COVID-19 |  |  |  |
| --- | --- | --- | --- | --- |
|  | Total causal effect | Direct effect | Indirect effect | Proportion mediated |
| IMD - 1 | 1.3 (0.88, 2.80) | 1.3 (1.00, 1.56) | 1.0 (0.72, 2.54) | 0.01 (-4.38, 1.65) |

The reference group was taken as deprivation quintile: IMD 5 (least deprived). Only IMD quintile pairwise comparisons with disproportionate total causal effect were included in the mediation analysis (IMD1)
Exposure: deprivation; Pre-exposure confounders: age group, sex, region, ethnic group; Mediator: comorbidities

### COVID-19

#### Total causal effects

No ethnic group had disproportionately high odds of COVID-19 hospitalization (model 1, Table 3b). Four of the 15 ethnic groups had disproportionately low odds of COVID-19 hospitalization; the Black African group (OR 0.5, 95% CI: 0.44, 0.58), the Any other Black background group (OR 0.6, 95% CI: 0.45, 0.70), the Indian group (OR 0.6, 95% CI: 0.53, 0.65) and the Chinese group (OR 0.4, 95% CI: 0.32, 0.55). Only the most deprived level (IMD1) had disproportionately high odds compared with the least deprived, with odds 1.5 (95% CI: 1.43, 1.55) (model 2, Table 4b), a smaller difference than observed for influenza.

### Mediation analysis

Ethnic group mediation was not conducted due to the absence of disproportionate total effects. For IMD1, the indirect effect mediated by vaccination uptake, conditioning on comorbidities, was significant (Figure 3, Table 6b) (OR 1.2, 95% CI: 1.01, 1.20). The estimated proportion mediated was high (97%) but unstable, with wide confidence intervals, limiting interpretability. No significant indirect effects were observed for comorbidities alone (Table 6d). Overall, after accounting for vaccination uptake and comorbidities, no meaningful residual deprivation inequality in COVID-19 hospitalization remained.

### Model checks

For total causal effect models with ethnic group as the exposure, inclusion of age, sex, and region as competing exposures reduced both the Akaike Information Criterion (AIC) and Bayesian Information Criterion (BIC) relative to unadjusted models for influenza and COVID-19 (Supplementary Table 6). This indicated improved model fit, and these adjusted models were therefore retained as the primary total causal effect specifications.

We assessed whether including mediator-exposure interaction terms in the outcome models improved goodness of fit. For influenza, when ethnic group was the exposure, and deprivation the mediator, inclusion of an interaction term increased both AIC and BIC (Supplementary Table 7, Table 1), indicating poorer model fit. Mediation by ethnic group was not conducted for COVID-19, therefore interaction testing was not applicable for this specification.

When deprivation was the exposure and vaccination uptake the mediator, inclusion of a mediator-exposure interaction produced mixed evidence. For influenza, the interaction decreased BIC but increased AIC (Supplementary Table 7, Table 2), suggesting that while the interaction captured some systematic structure in the data, it did not materially improve predictive performance [25]. In contrast, for COVID-19, both AIC and BIC decreased, indicating improved model fit. Mediation results including interaction terms for vaccination uptake were compared with primary analyses and showed consistent effect estimates and conclusions for both infections (Supplementary Table 8, Tables 1 – 2).

Variance inflation factors (VIF) for all covariates in the models (excluding interactions terms) were below five, indicating no evidence of problematic multicollinearity under standard thresholds [35, 36]. Interaction terms were excluded from VIF assessment, as elevated VIFs are expected in the presence of interactions.

### Sensitivity analyses: comorbidities

Because comorbidities form part of the eligibility criteria for vaccination, analyses restricted to the eligible cohort are subject to selection bias. Among individuals aged 17–64, eligibility is based solely on comorbidity status, whereas children aged 2–16 (influenza) and adults aged ≥65 (influenza and COVID-19) include individuals both with and without comorbidities due to age-based eligibility. As a result, in some age strata, comorbidity status is deterministically related to eligibly, leading to limited variation and structural dependence between age and comorbidity indicators. Consequently, estimates from the eligible cohort primarily describe how inequalities operate within this selected population, rather than the contribution of comorbidities in the general population.

To assess the impact of this selection, we repeated mediation analyses stratified by age group. For influenza, results for ages 2–16 and ≥65 were compared with the full eligible cohort analysis. For COVID-19, results for those aged ≥65 were compared with the full eligible cohort analysis. Mediation by ethnic group was not examined for COVID-19, due to the absence of disproportionate total effects.

For influenza, when ethnicity was the exposure, comorbidities were not a significant mediator among children aged 2–16 (Supplementary Table 14, Table 1b), consistent with results from the full eligible cohort analysis (Table 5). Among adults aged ≥65, however, comorbidities significantly mediated ethnic group differences, conditioning on deprivation, with indirect effects of 1.2 for both Pakistani and Bangladeshi groups, corresponding to proportions mediated of 33% and 29%, respectively. Deprivation remained an important mediator of ethnic group effects in both age strata (Supplementary Table 14, Table 1a).

When deprivation was the exposure, comorbidities were not a significant mediator of the deprivation-hospitalization association for either infection in age-stratified analyses, consistent with the full eligible cohort analysis (Supplementary Table 15, Tables 3–4).

However, for influenza, age stratification decreased evidence for mediation by vaccination uptake when conditioning on comorbidities, as indirect effects were no longer significant in these models (Supplementary 15, Table 1). In contrast, for COVID-19, among those aged ≥65, almost all the deprivation effect was mediated through vaccination uptake, conditioning on comorbidities (Supplementary Table 15, Table 2), whereas no residual deprivation effect was observed in the full eligible cohort analysis (Table 6).

We also repeated all mediation analyses for the whole population, where comorbidities were not subject to selection. Patterns of total ethnic and deprivation inequalities were similar to those observed in the eligible cohort, although effect sizes were marginally larger in some cases (Supplementary Tables 9–10). In the whole-population mediation analyses with ethnicity as the exposure (influenza only, Supplementary Table 11), deprivation remained important mediator. Unlike in the eligible cohort, comorbidities mediated part of the ethnic group effect for the Bangladeshi group (indirect effect OR = 1.1, 95% CI: 1.02, 1.18, proportion mediated 16%). For deprivation as the exposure, vaccination uptake mediated influenza inequalities for the most deprived group (IMD1), while for COVID-19, adjustment for vaccination uptake and comorbidities eliminated residual deprivation inequalities (Supplementary Table 13), consistent with eligible-cohort findings. Unlike eligible-cohort analyses, comorbidities alone were an important mediator of COVID-19 deprivation inequalities in the whole population (indirect effect OR = 1.2, 95% CI: 1.05, 1.37), proportion mediated 51%), in contrast to results from the eligible cohort.

Overall, within analyses restricted to the full eligible cohort, comorbidities did not appear to mediate ethnic or deprivation inequalities, partly because eligibility itself is determines by comorbidity status. Vaccination policy targets those most at risk, partly based on comorbidity status, which aims to mitigate inequalities between ethnic group and deprivation inequalities in the whole population. Within the eligible cohort, in age groups with variation in comorbidities, and in analyses of the whole population, comorbidities played a more substantial mediating role for selected groups and outcomes. Because whole-population analyses include additional causal pathways not related to vaccination eligibility, findings from the eligible cohort should not be extrapolated beyond this selected population.

## Discussion

In the vaccine-eligible cohort, inequalities in hospitalization were substantially greater for influenza than for COVID-19. For influenza, people from the Pakistani and Bangladeshi groups had disproportionately higher odds of hospitalization than the White British group. No ethnic groups had disproportionately higher odds of COVID-19 hospitalization, with several groups, including the Chinese and Black African groups, with lower odds than the White British group, which may reflect differences in infection exposure, social and occupational contact patterns, healthcare-seeking behaviours, and uptake of protective health behaviours. Deprivation-based inequalities were evident for both types of infection but were larger for influenza.

The comparatively lower risk of COVID-19 hospitalization for ethnic minority groups found in this study is in stark contrast to early stages of the pandemic (2020 - 2021) when most ethnic groups had significantly worse outcomes compared to the White British group. Given widespread evidence of current ethnic group inequalities in respiratory disease [26], our finding is perhaps surprising. However, it is congruent with other research: the ONS found no evidence of ethnic minority groups having higher COVID-19 mortality compared to the White British group during the Omicron period (Jan - Dec 2022) [27]. Historically, increased risk of infection was the main driver of disproportionate clinical outcomes for COVID-19 in ethnic minority groups, with smaller differences observed for subsequent outcomes, including hospitalization [6]. Ethnic minority groups may have acquired higher levels of immunity to SARS-CoV-2 because of their greater likelihood of previous infection, which, in turn, may have contributed to the attenuation of ethnic group inequalities in COVID-19 hospitalization in more recent periods [28].

Deprivation was an important mediator of ethnic group inequalities for influenza. For the Pakistani and Bangladeshi groups, approximately 26% to 41% of the excess odds of hospitalization were mediated through differences in deprivation. These estimates will include the unmeasured variables in our DAG (Figure 1), and differences in vaccination uptake – our approach was unable to decompose these for ethnic group. Other unmeasured variables linked to deprivation and contributing to the effect may also include persistent differences in occupational exposure to infection [29–31], housing and neighbourhood conditions, including exposure to air pollution [32, 33], and psychosocial and sociocultural determinants for health-related behaviours [34]. Lifestyle factors, such as smoking [35], are closely linked to deprivation, further increase vulnerability to severe respiratory infection. Many of these determinants are shaped by wider social and economic policies and deserve further research and evaluation. Tackling deprivation would offer widespread health benefits but is inherently more challenging than improving vaccination coverage.

Vaccination uptake was an important mediator in deprivation inequalities for influenza and COVID-19 hospitalization, after accounting for differences between deprivation levels in age, sex, region, ethnic group and comorbidities. For influenza, differences in vaccination uptake accounted for between 20% - 30% of the excess hospitalization risk among people living in the most deprived areas (IMD1 and IMD2). For COVID-19, there was no residual difference between deprivation levels after including vaccination uptake and comorbidities. As a modifiable factor, vaccination uptake represents a clear and actionable target for public health interventions to reduce inequalities for both respiratory infections – if vaccination uptake was increased across ethnic groups and deprivation levels with low coverage, hospitalization rates would reduce and be more similar to the White British group, and the least deprived areas.

Within the eligible cohort, comorbidities did not play a significant mediating role in deprivation inequalities, nor in ethnic group inequalities, after conditioning on deprivation. However, as vaccine eligibility in some age groups was based on comorbidity status, the lack of effect may be explained by selection bias. Comorbidities remain an important determinant of hospitalization overall, and in mediating inequalities for certain groups, as shown in whole-population analyses and age-stratified analyses of the eligible cohort (Supplementary Tables 9-15). Whilst the prevalence of comorbidities could be reduced, this would require long-term changes across the health system, so they are less amenable to policy action than vaccination uptake – apart from their existing application in prioritizing vaccination.

Several limitations should be acknowledged. The results depend on the assumptions encoded in the DAG and mediation models. For ethnic group, we only considered the proportion of the effect mediated by deprivation and comorbidities; the remaining direct effect could be decomposed further into the effect due to vaccination uptake or other unmeasured factors. For deprivation, we only considered the role of comorbidities, and then vaccination uptake, conditioned on comorbidities. Future work should explore more complex mediation structures involving multiple intermediate confounders. We also made simplifying assumptions about the variables included in the DAG, and the temporal order of variables, for instance, that region precedes an individual’s deprivation status. There are likely to be more complex interactions over time and additional unmeasured factors not included here. This analysis did not examine inequalities in infection risk, healthcare access, or care experiences, for instance due to racism or discrimination [36], nor outcomes following hospitalization. Findings reflect hospitalization risk among infected individuals within the vaccine eligible cohort during a 2023-24 in England and may not generalize to other contexts or time periods.

Improving vaccination uptake in groups with low coverage would likely reduce hospitalization inequalities. Lessons from the COVID-19 pandemic highlight the importance of accessible and inclusive communications which address vaccine concerns, and the use of trusted community and healthcare networks to provide accurate information [37]. Tailored, locally co-designed solutions, alongside strengthened core national vaccination offers in General Practice and to school-age children, must remain a policy priority [38]. However, reducing ethnic and socioeconomic inequalities in respiratory infections requires wider action on the underlying social determinants of health, particularly poverty, alongside efforts to improve vaccination uptake.

## Data Availability

The UK Health Security Agency (UKHSA) operates a robust governance process for applying to access protected data that considers
1. The benefits and risks of how the data will be used
2. Compliance with policy, regulatory and ethical obligations
3. Data minimization
4. How the confidentiality, integrity, and availability will be maintained
5. Retention, archival, and disposal requirements
6. Best practice for protecting data, including the application of privacy by design and by default, emerging privacy conserving technologies and contractual controls
Access to protected data is always strictly controlled using legally binding data sharing contracts. UKHSA welcomes data applications from organizations looking to use protected data for public health purposes. To request an application pack or discuss a request for UKHSA data you would like to submit, contact.

## Acknowledgements

We thank NHS England for providing the data used in this analysis. We are grateful for the contributions and review from UKHSA colleagues, including Steve Parratt and the following teams: Public Health Analysis, Health Equity and Inclusion Health, Statistics, Infectious Disease Modelling, Immunisations and Vaccine Preventable Diseases. We would also like to thank the anonymous reviewers for their helpful suggestions which greatly improved the paper.

## Data availability statement

The UK Health Security Agency (UKHSA) operates a robust governance process for applying to access protected data that considers:

- the benefits and risks of how the data will be used
- compliance with policy, regulatory and ethical obligations
- data minimization
- how the confidentiality, integrity, and availability will be maintained
- retention, archival, and disposal requirements
- best practice for protecting data, including the application of ‘privacy by design and by default’, emerging privacy conserving technologies and contractual controls

Access to protected data is always strictly controlled using legally binding data sharing contracts. UKHSA welcomes data applications from organizations looking to use protected data for public health purposes. To request an application pack or discuss a request for UKHSA data you would like to submit, contact.

## References

1. UK Health Security Agency, *Health equity for health security strategy 2023 to* 2026. 2024, UK Health Security Agency: https://www.gov.uk/government/publications/ukhsa-health-equity-for-health-security-strategy-2023-to-2026.

2. UK Health Security Agency, Infectious diseases impacting England: 2025 report. 2025: https://www.gov.uk/government/publications/infectious-diseases-impacting-england-2025-report.

3. Zhao, H., et al., Ethnicity, deprivation and mortality due to 2009 pandemic influenza A(H1N1) in England during the 2009/2010 pandemic and the first post-pandemic season. Epidemiology and Infection, 2015. 143(16).

4. Chandrasekhar, R., et al., Social determinants of influenza hospitalization in the United States. Influenza and other respiratory viruses, 2017.

5. Davidson, J., et al., Ethnic differences in the incidence of clinically diagnosed influenza: an England population-based cohort study 2008-2018. Wellcome Open Research, 2021.

6. Irizar, P., et al., Ethnic inequalities in COVID-19 infection, hospitalisation, intensive care admission, and death: a global systematic review and meta-analysis of over 200 million study participants. eClinicalMedicine, 2023. 57.

7. UK Health Security Agency, *Inequalities in emergency hospital admission rates for influenza and COVID-19, England: September 2022 to February* 2023. 2023: https://www.gov.uk/government/publications/covid-19-and-flu-inequalities-in-emergency-hospital-admission-rates.

8. UK Health Security Agency, National Influenza and COVID-19 surveillance report: week 3 report (up to week 2 2024 data). 2024, UK Health Security Agency, Immunisations and Vaccine-Preventable Diseases.

9. UK Health Security Agency, Seasonal influenza vaccine uptake in GP patients in England: winter season 2023 to 2024. 2024, UK Health Security Agency, Immunisations and Vaccine-Preventable Diseases.

10. Public Health England, *COVID-19: review of disparities in risks and outcomes*. 2020, PHE publications gateway number: GW-1447: https://www.gov.uk/government/publications/covid-19-review-of-disparities-in-risks-and-outcomes.

11. Ramsay, M.E., Immunisation against infectious disease, U.H.S. Agency, Editor. 2024: Immunisation against infectious disease.

12. Bosworth, M., et al., *Inequalities in mortality involving common physical health conditions, England: 21 March 2021 to 31 January* 2023, H.D. Office for National Statistics, Editor. 2023: https://www.ons.gov.uk/releases/sociodemographicandgeographicaldifferencesin ratesofhospitaladmissionandmortalityforcommonphysicalhealthconditionsengland 21march2021to31january2023.

13. Petersen, J., J. Kandt, and P.A. Longley, Ethnic inequalities in hospital admissions in England: an observational study. BMC Public Health, 2021.

14. The Health Foundation. Inequalities in diagnosed health conditions by ethnicity. 2025 [cited 2025 27/03/2025].

15. Hayanga, B., M. Stafford, and L. Bécares, Ethnic inequalities in multiple long-term health conditions in the United Kingdom: a systematic review and narrative synthesis. BMC Public Health, 2023. 23.

16. Office for National Statistics, *Updating ethnic contrasts in deaths involving the coronavirus (COVID-19), England: 8 December 2020 to 1 December* 2021. 2022.

17. Herbert, A., et al., Data resource profile: Hospital episode statistics admitted patient care (HES APC), I.J.o. Epidemiology, Editor. 2017.

18. Tessier, E., Edelstein, M., Tsang, C., Krisebom, F., Gower, C., Campbell, C., Ramsay, M., White, J., Andrews, N., Lopez-Bernal, J. & Stowe, J., Monitoring the COVID-19 immunisation programme through a national immunisation Management system – England’s experience. International Journal of Medical Informatics, 2023. 170(104974).

19. Tennant, P.W.G., et al., Use of directed acyclic graphs (DAGs) to identify confounders in applied health research: review and recommendations. International Journal of Epidemiology, 2021. 50(2): p. 620–632.

20. Office for National Statistics, *Ethnic group by age and sex, England and Wales: Census* 2021, Office for National Statistics, Editor. 2023: Ons.gov.uk.

21. Valeri, L.V., T., Mediation analysis allowing for exposure–mediator interactions and causal interpretation: theoretical assumptions and implementation with SAS and SPSS macros. Psychological methods, 2013. 18(2): p. 137–150.

22. Shi, B., et al., CMAverse a suite of functions for reproducible causal mediation analyses. Epidemiology, 2021. 32(5).

23. Robins, J., A new approach to causal inference in mortality studies with a sustained exposure period—application to control of the healthy worker survivor effect. Mathematical Modelling, 1986. 7.

24. Race Equality Unit, Using relative likelihoods to compare ethnic disparities. 2020: gov.uk.

25. Shmueli, G., “To Explain or to Predict?*.”.* Statistical Science, 2010. 25.

26. UK Health Security Agency, Health inequalities in health protection report. 2025: https://www.gov.uk/government/publications/health-inequalities-in-health-protection-report.

27. Office for National Statistics, *Updating ethnic and religious contrasts in deaths involving the coronavirus (COVID-19), England: 24 January 2020 to 23 November* 2022. 2023, Office for National Statistics: https://www.ons.gov.uk/peoplepopulationandcommunity/birthsdeathsandmarriages/deaths/articles/updatingethniccontrastsindeathsinvolvingthecoronaviruscovid19englandandwales/24january2020to23november2022.

28. Pan, D., et al., Are clinical outcomes from COVID-19 improving in ethnic minority groups? eClinicalMedicine, 2023. 61.

29. Industrial Injuries Advisory Council, *COVID-19 and occupational impacts*. 2022, Industrial Injuries Advisory Council, IIAC: https://www.gov.uk/government/publications/covid-19-and-occupational-impacts/covid-19-and-occupational-impacts#summary.

30. Pearce, N., et al., Occupational differences in COVID-19 incidence, severity, and mortality in the United Kingdom: Available data and framework for analyses. Wellcome Open Research, 2023.

31. Rhodes, S., et al., Occupational differences in SARS-CoV-2 infection: analysis of the UK ONS COVID-19 infection survey. Journal of epidemiology and community health, 2022. 76: p. 841–846.

32. Balogun, B., F. Rankl, and W. Wilson, Health inequalities: cold or damp homes. 2023, Library, House of Commons: Library, House of Commons.

33. Department for Levelling Up Housing and Communities, *Housing with damp problems*. 2020, gov.uk: https://www.ethnicity-facts-figures.service.gov.uk/housing/housing-conditions/housing-with-damp-problems/latest/.

34. Flowers, P., et al., Understanding pandemic influenza behaviour: An exploratory biopsychosocial study. Journal of Health Psychology, 2016.

35. Baskaran, V., W.S. Lim, and T.M. McKeever, Effects of tobacco smoking on recurrent hospitalisation with pneumonia: a population-based cohort study. British Medical Journal, 2021. 77: p. 82–85.

36. National Health Service England, Improving access for all: reducing inequalities in access to general practice services. 2018, National Health Service England, NHS England: https://www.england.nhs.uk/wp-content/uploads/2017/07/inequalities-resource-sep-2018.pdf.

37. Kamal, A., A. Hodgson, and J.M. Pearce, A Rapid Systematic Review of Factors Influencing COVID-19 Vaccination Uptake in Minority Ethnic Groups in the UK. Vaccines, 2021. 9(10)(1121).

38. UK Health Security Agency, *National Immunisation Programme Health Equity Audit* 2025. 2026: https://www.gov.uk/government/publications/national-immunisation-programme-health-equity-audit-2025/national-immunisation-programme-health-equity-audit-2025#actions-required-to-reverse-inequalities.

